# HIV Risk Behavior Profiles Among Men Who Have Sex with Men Interested in Donating Blood: The Assessing Donor Variability and New Concepts in Eligibility (ADVANCE) Study

**DOI:** 10.1101/2023.04.08.23288320

**Authors:** Brian Custer, Barbee Whitaker, Lance Pollack, Renata Buccheri, Roberta Bruhn, Lauren Crowder, Susan L. Stramer, Rita Reik, Suchitra Pandey, Mars Stone, Clara Di Germanio, Kate Buchacz, Anne Eder, Yun Lu, Richard Forshee, Steven Anderson, Peter Marks

## Abstract

**Importance:** Blood donor selection policies should be evidence-based. Individual risk assessment allows potential donors to be evaluated based on their own behaviors.

**Objective:** The Assessing Donor Variability and New Concepts in Eligibility (ADVANCE) study examined behavioral and biomarkers of HIV risk in sexually active men who have sex with men (MSM) to estimate the proportion of the study population who would not be deferred for higher risk HIV sexual behaviors and might be eligible to donate.

**Design:** A cross-sectional assessment of sexually active MSM interested in blood donation. Setting: An 8-city study of MSM aged 18 – 39 years assigned male sex at birth.

**Interventions or Exposures:** Participants completed surveys during 2 study visits to define eligibility, self-reported sexual and HIV prevention behaviors. Blood was drawn at study visit 1 and tested for HIV and the presence of tenofovir, 1 of the drugs in oral HIV pre-exposure prophylaxis (PrEP).

**Main Outcomes and Measures:** Associations between HIV infection status or HIV PrEP use and self-reported HIV risk behaviors, including number of male sex partners, new partners, and anal sex.

**Results:** Among 1788 screened MSM, 1593 were eligible and 1566 completed the visit 1 HIV risk questionnaire and blood draw. A median of 22 days later, 1197 completed the visit 2 follow-up questionnaire. Four individuals tested HIV positive (0.25%). Among HIV-negative participants, 789 (50.4%) reported no PrEP use in the past 3 months. The number of sex partners in the past 3 months was significantly higher among PrEP users versus non-users, as was the number reporting a new male sex partner in the same period. Among HIV-negative, non-PrEP using participants, 66.2% reported only 1 sexual partner or no anal sex and 69% reported no new sexual partners or no anal sex with a new partner in the past 3 months.

**Conclusion and Relevance:** Among sexually active MSM, there are subgroups who self-report no new sexual partners and only 1 sexual partner within the past 3 months. These individuals are likely at lower risk of HIV infection than other MSM and would meet proposed individual risk assessment criteria for blood donation in the U.S.

**Key Points:** *Question:* Can a set of blood donor screening questions based on individual behaviors identify a population of lower HIV risk men who have sex with men (MSM) who wish to donate blood?

*Findings:* In this cross-sectional study of 1566 enrolled MSM, among 1562 persons without HIV, 789 (50.4%) were not taking pre-exposure prophylaxis (PrEP). Of those not taking PrEP, 66.2% reported only 1 sexual partner or no anal sex and 69% reported no new sexual partners or no anal sex with a new partner in the past 3 months.

*Meaning:* Potential blood donor history questions were able to identify sexually active MSM with lower-risk sexual behaviors who may be eligible to donate blood.

## Introduction

Blood donor deferral related to HIV began in 1983 when men who have sex with men (MSM) along with other groups with higher risk of HIV infection^1^ were deferred indefinitely from donation. In the subsequent 40 years, blood donation testing has evolved. Now, all blood donations are screened using sensitive HIV serology (since 1985) and nucleic acid tests (NAT, since 1999).^2^ In 2015, given performance of testing technology and results from studies, such as the Retrovirus Epidemiology Donor Studies^3-5^ the U.S. Food and Drug Administration (FDA) revised guidance on blood donor eligibility allowing donation by MSM following a 12-month deferral period since the last oral or anal sex with a man.^6^ In 2020, given results from the Transfusion Transmissible Infections Monitoring System (TTIMS) following the change from a lifetime deferral to a time-based deferral for MSM^7,8^ and the negative impact of the SARS-CoV-2/COVID-19 pandemic on the U.S. blood supply, FDA again revised blood donor eligibility for MSM to 3 months since last sexual encounter. Three months amply covers the NAT window period during which the virus cannot be detected by the test but may be present and could infect a transfusion recipient.^9^ Most U.S. blood collection organizations implemented the new deferral criteria in mid-2020.

Compared to other groups, MSM are at higher risk for HIV and account for more than 70% of new HIV diagnoses in the U.S. and about 80% of those among men.^10^ HIV risk in the MSM population is associated with condomless anal sex and the per sex act risk of infection increases with the number of sexual partners.^11^ As part of Ending the HIV Epidemic (EHE) by 2030, the U.S. seeks to reduce inequities in access to HIV treatment and prevention and to increase access to pre-exposure prophylaxis (PrEP).^12^ PrEP, in combination with safer sex practices, is indicated to reduce the risk of sexually acquired HIV in adults at high risk.^13,14^ PrEP use, the behavioral characteristics of PrEP users, and differences in oral and injected PrEP^15,16^ have implications for the safety of the blood supply due to the possibility of breakthrough HIV infection and window period donations that could infect a transfusion recipient.^17,18^ PrEP use is a blood donation deferral criterion because of the concern that a PrEP-breakthrough infection would be undetected by screening assays.^19^ Consequently, complex U.S. blood donation policy changes must be evaluated within the context of contemporary HIV prevention strategies and donor selection procedures.

In January 2023, FDA released draft guidance proposing to move from time-based deferrals to individual risk assessments for all prospective blood donors.^20^ Recently, the United Kingdom and Canada have transitioned from 3-month deferral since last sex for MSM to individual risk questions, focusing on the use of PrEP and post-exposure prophylaxis, the number of sexual partners, and specific sexual behaviors.^21-24^ The FDA recommendations approximately mirror these donor selection approaches. The Assessing Donor Variability and New Concepts in Eligibility (ADVANCE) study was designed to assess among U.S. MSM interested in donating blood whether simple questions differentiate those with lower and higher risk of HIV exposure. We report findings from the ADVANCE study, focused on differences in self-reported behaviors among MSM.

## Methods

### Study Design

The ADVANCE study is a cross-sectional behavioral and biomarker assessment of sexually active MSM conducted in 8 U.S. metropolitan areas (Atlanta, GA; Los Angeles, CA; Memphis, TN; Miami, FL; New Orleans/Baton Rouge, LA; Orlando, FL; San Francisco, CA; and Washington DC) between December 2020 and November 2022. This study was conducted by 3 large blood collection organizations (Vitalant, OneBlood, and the American Red Cross) with assistance from Stanford Blood Center, LGBTQ+ organizations in each city, the University of California San Francisco Center for AIDS Prevention Studies, and the FDA. The study protocol was approved by the Institutional Review Board, Advarra (Columbia, MD), under protocol number 00043278. A Certificate of Confidentiality was obtained from the Division of Inspections and Surveillance, Office of Compliance and Biologics Quality, Center for Biologics Evaluation and Research, FDA, protecting the participants from release of any study information except when required by law.

### Study Population

Sexually active MSM were recruited to participate in the study. Individuals were required to be biologically male at birth, reside within the designated geographic area (by zip code), and between 18 to 30 years old to include the MSM demographic at highest risk of new HIV infection. Age eligibility was expanded on May 10, 2021, to 18 to 39 years to help increase enrollment. Individuals were excluded who: did not report having oral or anal sex with a male within the past 3 months; were ever diagnosed with HIV; reported injection drug use, exchanging money or goods for sex, or had been diagnosed with a sexually transmitted infection (syphilis, gonorrhea, chlamydia) in the past 3 months [all these criteria, except men not having sex with another man, are exclusions from donation]; and/or were not able to complete the study documents in English or Spanish. When data collection began, follow-up questionnaire eligibility was restricted to participants who tested HIV-positive or tested HIV-negative and PrEP-reactive. In May 2021, follow-up questionnaire eligibility was expanded to include participants who tested HIV-negative and PrEP non-reactive or inconclusive but who reported PrEP use on the HIV risk questionnaire, and in July 2021 to all participants, due to the relevance of follow-up survey responses for study objectives.

### Participant Recruitment

Participant recruitment was supported using multiple outreach methods. In partnership with LGBTQ+ community organizations, the ADVANCE study was promoted using printed material, by direct email invitations, and via social media. Recruitment was also conducted at in-person events (e.g., Pride events, concerts) after relaxation of COVID-19 restrictions. All interested persons were referred to <u>the study website</u> to learn more and to schedule an in-person eligibility visit. Participants were compensated a maximum of $85.

### Study Visits

Participants completed eligibility screening at their first study visit. Eligible and consented participants completed a set of HIV risk questions focused on different time intervals (past 1, 3, and 12 months) inquiring about number of sex partners, sexual behaviors, HIV-status of sex partners, and use of PrEP. PrEP use was ascertained by questions covering 3 intervals, i.e., Have you taken pre-exposure prophylaxis (PrEP) during the past month [3 months], [12 months]? Responses were entered in an electronic informed consent and data capture system (Medrio, Inc. San Francisco, CA) using tablet computers provided by the study. Participants then provided a whole blood phlebotomy sample totaling <30mL for testing for HIV and tenofovir testing.

During the second study visit, median of 22 days after the first, participants were informed of their HIV and PrEP results and participants with HIV (hereafter referred to as HIV-positive) were referred to local HIV treatment clinics. Participants were consented for additional study activities and then completed the follow-up questionnaire that included questions from the CDC National HIV Behavioral Surveillance (NHBS) survey, 2021-2022 MSM cycle.^25^ Content was designed to elicit additional behavior information (e.g., number and sex/gender of sexual partners, sexual behaviors, HIV prevention including types of PrEP and condom use) as well as motivations and interest in blood donation.

### HIV RNA and Antibody Testing

Creative Testing Solutions (Tempe, AZ) conducted NAT to detect HIV RNA using the Ultrio Elite assay on the Panther System (Grifols Diagnostics, San Diego, CA) and for antibodies using the HIV-1/HIV-2 PLUS O enzyme immunoassay (Bio-Rad, Hercules, CA).

### HIV Limiting Antigen and Tenofovir Testing

Limiting Antigen (LAg)-Avidity enzyme immunoassay testing (Sedia Biosciences®, Portland, OR) was conducted at VRI and was used to define the infection as recently acquired or long-standing among those who tested HIV NAT and serology reactive.^26^

Tenofovir is 1 of the 2 antiretroviral drugs in oral PrEP medications. We used a plate-based enzyme linked immunosorbent assay (ELISA, from OraSure®, Bethlehem, PA) with whole blood as the specimen input type to assess biomarkers of PrEP use. Additional information on the study testing is provided in the supplement.

### Data Analysis

To compare behavioral profiles, primary analysis focused on frequencies of HIV risk questionnaire responses stratified by HIV-status and self-reported PrEP use. Chi-square statistics tested for significant statistical association (*p* ≤ 0.05). Secondary analyses assessed the proportion of participants eligible for blood donation as recommended by FDA^20^ using a sequence of questions beginning with PrEP use in the past 3 months followed by 2 sets of questions first pertaining to the number of sex partners in the past 3 months and a question about anal sex if there has been more than 1 partner, and second about any new sex partners in the past 3 months and a question about anal sex if there has been a new partner. We used HIV risk questionnaire responses to estimate the number of donation-eligible participants for the “number of sex partners” questions and responses from the HIV risk and the follow-up questionnaires to estimate the number of donation-eligible participants for the “new partner” questions.

## Results

Participant enrollment and follow-up study visits occurred from December 7, 2020, to November 9, 2022. Of 1781 participants screened for eligibility, 1588 (89.2%) were eligible and consented to participate (Table 1). Common reasons for ineligibility were age or reporting a recent sexually transmitted infection. Of those eligible, 1566 (98.6%) completed the HIV risk questionnaire and blood draw of whom 1561 (99.7%) were notified of their test results. Accounting for shifting inclusion criteria over time, 1473 (94.4%) were eligible for the follow-up survey, 1200 (81.5%) consented, and 1197 (99.83%) completed the survey. Eligible and ineligible participants differed significantly (p<0.05) by demographic characteristics, except for age group, ethnicity, marital status, and previous blood donation (Table 2).

**Table 1.** ADVANCE Study Eligibility and Completion Rates

| **Study Participation Category** | **N** | **%**<br>**(of preceding row)** |
| --- | --- | --- |
| Total Screened | 1781 |  |
| Eligible for ADVANCE Study | 1593 | 89.4 |
| Consented for ADVANCE Study | 1588 | 99.7 |
| Completed HIV Risk Questionnaire | 1588 | 100 |
| Successful Blood Draw | 1566 | 98.6 |
| Notified of Blood Test Results | 1561 | 99.7 |
| Eligible for Follow-up Questionnaire* | 1473 | 94.4 |
| Consented for Follow-up Questionnaire | 1200 | 81.5 |
| Completed Follow-up Questionnaire | 1197 | 99.8 |
| Ineligible | 188 | 10.6** |
| Ineligibility reason*** |  | (% of ineligible) |
| Not biological male | 15<sup>a</sup> | 7.9 |
| <18 or >39 years of age | 48<sup>b</sup> | 25.5 |
| Zip code ineligible (out of area) | 27 | 14.4 |
| No oral or anal sex with a male past 3 months | 19 | 10.1 |
| Exchanging money or goods for sex past 3 months | 11 | 5.6 |
| IDU past 3 months | 1 | <1 |
| Self-reported HIV-positive | 15 | 8.0 |
| Positive for syphilis past 3 months | 4 | 2.1 |
| Positive for gonorrhea past 3 months | 47 | 25.0 |
| Positive for chlamydia past 3 months | 30 | 16.0 |
| Discontinued eligibility survey | 2<sup>c</sup> | 1.1 |

\* When data collection started on December 7, 2020, follow-up eligibility was restricted to participants who tested HIV-positive or tested HIV-negative and PrEP-reactive. Starting on May 10, 2021, participants who tested HIV-negative and PrEP negative/inconclusive but who reported any PrEP use in the HIV risk questionnaire became eligible for a follow-up survey. Starting on July 6, 2021, participants who tested HIV-negative and PrEP-negative/-inconclusive and did not report PrEP use in the HIV risk questionnaire also became eligible.
\*\*Of all presenting participants.
\*\*\*Respondents may have reported more than one reason for ineligibility; 159 (84.6%) were ineligible for 1 reason, 27 (14.4%) for 2 reasons, and 2 (1.1%) for 3 reasons.
<sup>a</sup> Includes 1 case who declined to answer the question (and was otherwise eligible).
<sup>b</sup> The number of age-ineligible participants (n=48) exceeds what is recorded in Table 2 for <18 and 40+ (n=39), because the upper limit of age eligibility expanded from 30 to 39 on May 10, 2021.
<sup>c</sup> Both participants were males within the eligible age range.

**Table 2.** Demographic and Social Characteristics of Eligible and Ineligible Respondents

| Variable | Category | Total | Eligible | Ineligible | p-value<sup>a</sup> |
| --- | --- | --- | --- | --- | --- |
|  |  | N=1781 (%) | N=1593 (%) | N=188 (%) |  |
| Site<sup>b</sup> | Atlanta | 179 (100) | 169 (94.4) | 10 (5.6) | <0.001 |
|  | Los Angeles | 345 (100) | 327 (94.8) | 18 (5.2) |  |
|  | Memphis | 59 (100) | 53 (89.8) | 6 (10.2) |  |
|  | Miami | 131 (100) | 117 (89.3) | 14 (10.7) |  |
|  | New Orleans/Baton Rouge | 108 (100) | 94 (87.0) | 14 (13.0) |  |
|  | Orlando | 193 (100) | 164 (85.0) | 29 (15.0) |  |
|  | San Francisco | 289 (100) | 234 (81.0) | 55 (19.0) |  |
|  | Washington, DC | 477 (100) | 435 (91.2) | 42 (8.8) |  |
| Gender Identity | Male | 1745 (98.0) | 1571 (98.6) | 174 (92.6) | <0.001 |
|  | Female<sup>c</sup> | 1 (0.1) | 0 | 1 (0.5) |  |
|  | Transgender<sup>c</sup> | 23 (1.3) | 11 (0.7) | 12 (6.4) |  |
|  | Prefer not to answer<sup>d</sup> | 12 (0.7) | 11 (0.7) | 1 (0.5) |  |
| Age | <18<sup>d</sup> | 1 (0.1) | 0 | 1 (0.5) | 0.724 |
|  | 18-24 | 250 (14.0) | 224 (14.1) | 26 (13.8) |  |
|  | 25-29 | 552 (31.0) | 506 (31.8) | 46 (24.5) |  |
|  | 30-34 | 527 (29.6) | 483 (30.3) | 44 (22.4) |  |
|  | 35-39 | 413 (23.2) | 380 (23.9) | 33 (17.6) |  |
|  | 40+<sup>d</sup> | 38 (2.1) | 0 | 38 (20.2) |  |
| Race | White/Caucasian | 1272 (71.4) | 1139 (71.5) | 133 (70.7) | 0.015 |
|  | Black/African American | 120 (6.7) | 99 (6.2) | 21 (11.2) |  |
|  | Asian/Pacific Islander | 127 (7.1) | 120 (7.5) | 7 (3.7) |  |
|  | Native American<sup>e</sup> | 11 (0.6) | 8 (0.5) | 3 (1.6) |  |
|  | More than 1 race | 149 (8.4) | 138 (8.7) | 11 (5.9) |  |
|  | Other | 91 (5.1) | 80 (5.0) | 11 (5.9) |  |
|  | Prefer not to answer<sup>d</sup> | 11 (0.6) | 9 (0.6) | 2 (1.1) |  |
| Ethnicity | Hispanic | 336 (18.9) | 298 (18.7) | 38 (20.2) | 0.621 |
|  | Non-Hispanic | 1442 (81.0) | 1293 (81.2) | 149 (79.3) |  |
|  | Prefer not to answer<sup>d</sup> | 3 (0.2) | 2 (0.1) | 1 (0.5) |  |
| Marital Status | Married/Domestic partner | 461 (25.9) | 409 (25.7) | 52 (27.7) | 0.859 |
|  | Separated/Divorced | 66 (3.7) | 58 (3.6) | 8 (4.3) |  |
|  | Widowed<sup>f</sup> | 3 (0.2) | 1 (0.1) | 2 (1.1) |  |
|  | Never married | 1204 (67.6) | 1080 (67.8) | 124 (66.0) |  |
|  | Other | 45 (2.5) | 43 (2.7) | 2 (1.1) |  |
|  | Prefer not to answer<sup>d</sup> | 2 (0.1) | 2 (0.1) | 0 |  |
| Education | Never been to school<sup>g</sup> | 0 | 0 | 0 | 0.029 |
|  | Completed grade school<sup>g</sup> | 2 (0.1) | 1 (0.1) | 1 (0.5) |  |
|  | Some high school<sup>g</sup> | 3 (0.1) | 2 (0.1) | 1 (0.5) |  |
|  | Completed high school | 49 (2.8) | 43 (2.7) | 6 (3.2) |  |
|  | Some college/tech school | 272 (15.3) | 231 (14.5) | 41 (21.8) |  |
|  | Completed college | 832 (46.7) | 755 (47.4) | 77 (41.0) |  |
|  | Completed graduate school | 621 (34.9) | 561 (35.2) | 60 (31.9) |  |
|  | Prefer not to answer<sup>d</sup> | 2 (0.1) | 0 | 2 (1.1) |  |

Table 2. Continued
| Variable | Category | Total | Eligible | Ineligible | p-value<sup>a</sup> |
| --- | --- | --- | --- | --- | --- |
| Employment Status | Full-time employment | 1355 (76.1) | 1234 (77.5) | 121 (64.4) | **<0.001** |
|  | Part-time employment | 147 (8.3) | 124 (7.8) | 23 (12.2) |  |
|  | Unemployed | 111 (6.2) | 87 (5.5) | 24 (12.8) |  |
|  | Currently in school | 151 (8.5) | 136 (8.5) | 15 (8.0) |  |
|  | Prefer not to answer<sup>d</sup> | 17 (1.0) | 12 (0.8) | 5 (2.7) |  |
| Annual Income | \$12,760 or less | 69 (3.9) | 53 (3.3) | 16 (8.5) | **0.032** |
| | \$12,761 - \$19,999 | 41 (2.3) | 35 (2.2) | 6 (3.2) | |
| | \$20,000 - \$29,999 | 80 (4.5) | 75 (4.7) | 5 (2.7) | |
| | \$30,000 - \$39,999 | 121 (6.8) | 106 (6.7) | 15 (8.0) | |
| | \$40,000 - \$49,999 | 136 (7.6) | 124 (7.8) | 12 (6.4) | |
| | \$50,000 - \$74,999 | 305 (17.1) | 271 (17.0) | 34 (18.1) | |
| | \$75,000 - \$99,999 | 244 (13.7) | 223 (14.0) | 21 (11.2) | |
| | \$100,000 or more | 698 (39.2) | 630 (39.5) | 68 (36.2) | |
|  | Don't know<sup>c</sup> | 43 (2.4) | 36 (2.3) | 7 (3.7) |  |
|  | Prefer not to answer<sup>c</sup> | 44 (2.5) | 40 (2.5) | 4 (2.1) |  |
| Last Donation | Never donated blood | 666 (37.4) | 599 (37.6) | 67 (35.6) | 0.971 |
|  | Over 5 years ago | 779 (43.7) | 696 (43.7) | 83 (44.1) |  |
|  | 1-5 years ago | 240 (13.5) | 214 (13.4) | 26 (13.8) |  |
|  | 3-12 months ago | 50 (2.8) | 47 (3.0) | 3 (1.6) |  |
|  | Within past 3 months<sup>h</sup> | 13 (0.7) | 10 (0.6) | 3 (1.6) |  |
|  | Prefer not to answer when<sup>d</sup> | 12 (0.7) | 10 (0.6) | 2 (1.1) |  |
|  | Prefer not to answer if ever<sup>d</sup> | 21 (1.2) | 17 (1.1) | 4 (2.1) |  |

Notes:
Los Angeles – enrollment sites in Los Angeles, Pasadena, and Culver City
San Francisco – enrollment sites in San Francisco, Oakland, and Palo Alto
Miami – enrollment sites in Miami and Fort Lauderdale
2 San Francisco participants discontinued the eligibility survey. They are included in the “Prefer not to answer” category for Education, Employment Status, and Annual Income and in the “Prefer not to answer if ever” for Last Donation In the “Ineligible” column.
<sup>a</sup> Chi-square statistic comparing eligible respondents to ineligible respondents
<sup>b</sup> Cell entries are row N and row percentage, all others are column N and column percentage
<sup>c</sup> Combined into “Other” category for calculation of chi-square statistic
<sup>d</sup> Excluded from calculation of chi-square statistic
<sup>e</sup> Combined with “Other” category for calculation of chi-square statistic

The study eligibility questionnaire asked if potential participants knew their HIV status. Among those screened for eligibility, 15 (0.84%) disclosed their HIV-positive status and were not eligible for the study. Four (0.25%) of the enrolled participants tested HIV-positive. The 4 had detectable HIV RNA and antibodies and were classified as not having recently acquired HIV infection by LAg avidity testing (Supplement Table 1). Tenofovir was detected for 1 of these participants. One participant who was unwilling to return was notified by certified mail; the others returned for in person results notification. At that time, 1 participant disclosed previous knowledge of his HIV infection, likely explaining the tenofovir result as part of antiretroviral therapy. Two of the HIV-positive participants completed the follow-up interview. Due to their small number, no demographic characteristics are reported for HIV-positive participants.

In the primary analysis of behavior and exposures that may be associated with HIV risk, we compared self-reported PrEP users to non-users. Among HIV-negative participants, 803 (51.7%) reported they did not use PrEP in the last month and 789/1552 (50.8%) reported no PrEP use in the last 3 months. In both the last month and last 3-month periods, the total number of sex partners was significantly different when comparing PrEP users and non-users, with 225 (30.0%) PrEP-users reporting 1 or no sex partner in the last month and 103 (13.5%) reporting 1 sex partner in the last 3 months, whereas 601 (74.8%) of non-PrEP users reported 1 or no sex partner in the last month and 492 (62.4%) reported 1 sex partner in the last 3 months (Table 3).

**Table 3.** Sexual Behavior with Male Sex Partners and PrEP Use Self-reported by MSM in the HIV Risk Questionnaire.

| Behavior | Category | Behavior in Past Month<sup>a</sup> |  |  |  | Behavior in Past 3 Months<sup>b</sup> |  |  |  |
| --- | --- | --- | --- | --- | --- | --- | --- | --- | --- |
|  |  | Tested HIV-negative |  |  | HIV-positive | Tested HIV-negative |  |  | HIV-positive |
|  |  | Used PrEP | No PrEP Use | p-value<sup>c</sup> |  | Used PrEP | No PrEP Use | p-value<sup>c</sup> |  |
|  |  | N=750 (%) | N=803 (%) |  |  | N=763 (%) | N=789 (%) |  |  |
| # male<br>partners | 0 | 21 (2.8) | 30 (3.7) | <0.001 | - | - | - | <0.001 | - |
|  | 1 | 204 (27.2) | 571 (71.1) |  | 2 (50) | 103 (13.5) | 492 (62.4) |  | 2 (50) |
|  | 2 | 147 (19.6) | 96 (12.0) |  | 1 (25) | 103 (13.5) | 105 (13.3) |  | - |
|  | 3-5 | 284 (37.9) | 84 (10.5) |  | - | 240 (31.5) | 136 (17.2) |  | 1 (25) |
|  | 6-10 | 74 (9.9) | 19 (2.4) |  | - | 208 (27.3) | 38 (4.8) |  | - |
|  | >10 | 20 (2.7) | 3 (0.4) |  | 1 (25) | 109 (14.3) | 17 (2.2) |  | 1 (25) |
|  | Missing data<sup>d</sup> | 0 | 0 |  | - | 0 | 1 (0.1) |  | - |
| Type of sex | No male sex partners<sup>e</sup> | 21 (2.8) | 30 (3.7) | <0.001 | - | - | - | <0.001 | - |
|  | No anal sex | 43 (5.7) | 122 (15.2) |  | - | 28 (3.7) | 83 (10.5) |  | - |
|  | Always used condoms | 57 (7.6) | 82 (10.2) |  | - | 49 (6.4) | 91 (11.5) |  | - |
|  | Sometimes used condoms | 224 (29.9) | 124 (15.4) |  | 3 (75) | 321 (42.1) | 166 (21.0) |  | 3 (75) |
|  | Never used condoms | 400 (53.3) | 442 (55.0) |  | 1 (25) | 363 (47.6) | 444 (56.3) |  | 1 (25) |
|  | Missing data<sup>d</sup> | 5 (0.7) | 3 (0.4) |  | - | 2 (0.3) | 5 (0.6) |  | - |
| Sex with<br>HIV-positive<br>partner | Yes | 70 (9.3) | 18 (2.2) | <0.001 | 1 (25) | 86 (11.3) | 22 (2.8) | <0.001 | 1 (25) |
|  | No | 659 (87.9) | 755 (94.0) |  | 3 (75) | 675 (88.5) | 766 (97.1) |  | 3 (75) |
|  | No male sex partners<sup>f</sup> | 21 (2.8) | 30 (3.7) |  | - | - | - |  | - |
|  | Missing data<sup>d</sup> | 0 | 0 |  | - | 2 (0.3) | 1 (0.1) |  | - |

<sup>a</sup> 4 HIV-negative participants declined to answer whether they took PrEP in the past month
<sup>b</sup> 5 HIV-negative participants declined to answer whether they took PrEP in the past 3 months
<sup>c</sup> Chi-square statistic comparing PrEP-users to non-users
<sup>d</sup> Excluded from calculation of chi-square statistic
<sup>e</sup> Combined with “No anal sex” category for calculation of chi-square statistic
<sup>f</sup> Combined with “No” category for calculation of chi-square statistic

Among those who completed the HIV risk questionnaire, 744 (96.4%) PrEP users had oral or anal sex in the past month and 772 (100%) in the past 3 months. For non-PrEP users 756 (96.8%) had oral or anal sex in the past month and 781 (100%) in the past 3 months (Table 4). Anal sex was reported by most HIV-negative respondents regardless of PrEP use or time interval, but the proportion was significantly higher among PrEP users compared to non-PrEP users. In each time period, both insertive and receptive anal sex were more common among PrEP users compared to non-PrEP users.

**Table 4.** Sexual Behavior and PrEP Use in Past 3 Months (Self-reported in HIV Risk Questionnaire) and Types of Sexual Partners and PrEP Use in Past 3 Months (Self-reported in Follow-up Questionnaire)

| Behavior | Most Recent Occurrence | Tested HIV-negative<sup>a</sup> |  |  | HIV-positive |
| --- | --- | --- | --- | --- | --- |
|  |  | Used PrEP Past 3 Months | No PrEP Use Past 3 Months | p-value<sup>b</sup> |  |
| HIV risk questionnaire responses |  | N=772 (%) | N=781 (%) |  | N=4 (%) |
| Oral/Anal Sex | Past month | 744 (96.4) | 758 (96.8) | 0.644 | 4 (100) |
|  | Past 3 months | 772 (100) | 781 (100) | -- | 4 (100) |
|  | Past 12 months | 772 (100) | 781 (100) | -- | 4 (100) |
| 2+ Partners | Past month | 534 (69.2) | 193 (24.7) | <0.001 | 2 (50) |
|  | Past 3 months | 668 (86.5) | 292 (37.4)<sup>c</sup> | <0.001 | 2 (50) |
|  | Past 12 months | 738 (95.6) | 401 (51.3) | <0.001 | 3 (75) |
| Anal Sex | Past month | 727 (94.2)<sup>c</sup> | 660 (84.5) | <0.001 | 4 (100) |
|  | Past 3 months | 751 (97.3) | 705 (90.3)<sup>c</sup> | <0.001 | 4 (100) |
|  | Past 12 months | 760 (98.4) | 740 (94.8)<sup>c</sup> | <0.001 | 4 (100) |
| Insertive Anal Sex | Past month | 543 (70.3)<sup>c</sup> | 467 (59.8) | <0.001 | 4 (100) |
|  | Past 3 months | 608 (78.8) | 538 (68.9)<sup>c</sup> | <0.001 | 4 (100) |
|  | Past 12 months | 671 (86.9) | 608 (77.8)<sup>c</sup> | <0.001 | 4 (100) |
| Receptive Anal Sex | Past month | 494 (64.0)<sup>c</sup> | 431 (55.2) | <0.001 | 3 (75) |
|  | Past 3 months | 559 (72.4)<sup>c</sup> | 508 (65.0)<sup>d</sup> | 0.002 | 3 (75) |
|  | Past 12 months | 632 (81.9)<sup>c</sup> | 578 (74.0)<sup>d</sup> | <0.001 | 4 (100) |
| Anal Sex Without Condoms | Past month | 636 (82.4)<sup>g</sup> | 552 (70.7)<sup>e</sup> | <0.001 | 4 (100) |
|  | Past 3 months | 694 (89.9)<sup>c</sup> | 617 (79.0)<sup>d</sup> | <0.001 | 4 (100) |
|  | Past 12 months | 718 (93.0) | 663 (84.9)<sup>c</sup> | <0.001 | 4 (100) |
| HIV-positive Partner | Past month | 71 (9.2) | 17 (2.2) | <0.001 | 1 (25) |
|  | Past 3 months | 92 (11.9)<sup>f</sup> | 22 (2.8)<sup>c</sup> | <0.001 | 1 (25) |
|  | Past 12 months | 151 (19.6)<sup>f</sup> | 27 (3.5)<sup>c</sup> | <0.001 | 1 (25) |
| Follow-up questionnaire responses |  | N=690 (%) | N=502 (%) |  | N=2 (%) |
| More Than 1 Partner Past 3 Months | Male | 569 (82.3)<sup>h</sup> | 178 (35.5)<sup>f</sup> | <0.001 | 1 (50) |
|  | Female | 6 (0.9)<sup>e</sup> | 13 (2.6)<sup>c</sup> | 0.020 | 0 |
|  | Transgender | 3 (0.4)<sup>k</sup> | 2 (0.4)<sup>h</sup> | 1.000 | 0 |
| New Partner Past 3 Months | Male | 541 (78.4)<sup>j</sup> | 167 (33.3)<sup>h</sup> | <0.001 | 1 (50) |
|  | Female | 7 (1.0)<sup>e</sup> | 14 (2.8)<sup>c</sup> | 0.022 | 0 |
|  | Transgender | 17 (2.5)<sup>l</sup> | 6 (1.2%)<sup>i</sup> | 0.115 | 0 |

Notes for HIV Risk Questionnaire Responses:
12-month data are adjusted for 3-month and 1-month responses, and 3-month data are adjusted for 1-month responses (including PrEP use)
<sup>a</sup> 4 HIV-negative participants declined to answer whether they took PrEP in the past 3 months
<sup>b</sup> For chi-square statistic comparing PrEP-users to non-users
<sup>c</sup> 1 case missing data
<sup>d</sup> 2 participants missing data
<sup>e</sup> 3 participants missing data
<sup>f</sup> 4 participants missing data
<sup>g</sup> 5 participants missing data
<sup>h</sup> 6 participants missing data

The follow-up questionnaire provides more specific information on sex in the last month and 3-month periods stratified by self-reported PrEP use in the last 3 months. Among PrEP users 569 (82.5%) reported more than 1 *male* sex partner in the past 3 months compared with 178 (35.5%) non-PrEP users (Table 4). The proportions reporting multiple *female* sex partners were small and significantly higher for non-PrEP users. Similarly, 541 (78.4%) PrEP users reported a new *male* sex partner in the past 3 months compared to 167 (33.3%) non-PrEP users. Few reported a new *female* sex partner, but they were significantly more common for non-PrEP users. No difference was evident regarding multiple or new *transgender* sex partners.

We estimated the proportion of the study population who would meet the proposed donor selection criteria in the FDA draft guidance for individual risk assessment questions. Each hierarchical analysis was restricted to HIV-negative participants. Among those who were not taking PrEP, 522 of 789 respondents (66.2%) reported fewer than 2 sex partners or not having anal sex with any partner in the past 3 months

(Figure 1a). A similar analysis of new sex partners in the last 3-months showed that among HIV-negative, non-PrEP users 352 of 510 (69.0%) did not have a new partner or did not have anal sex with a new partner in the past 3 months (Figure 1b).

**Figure 1.**
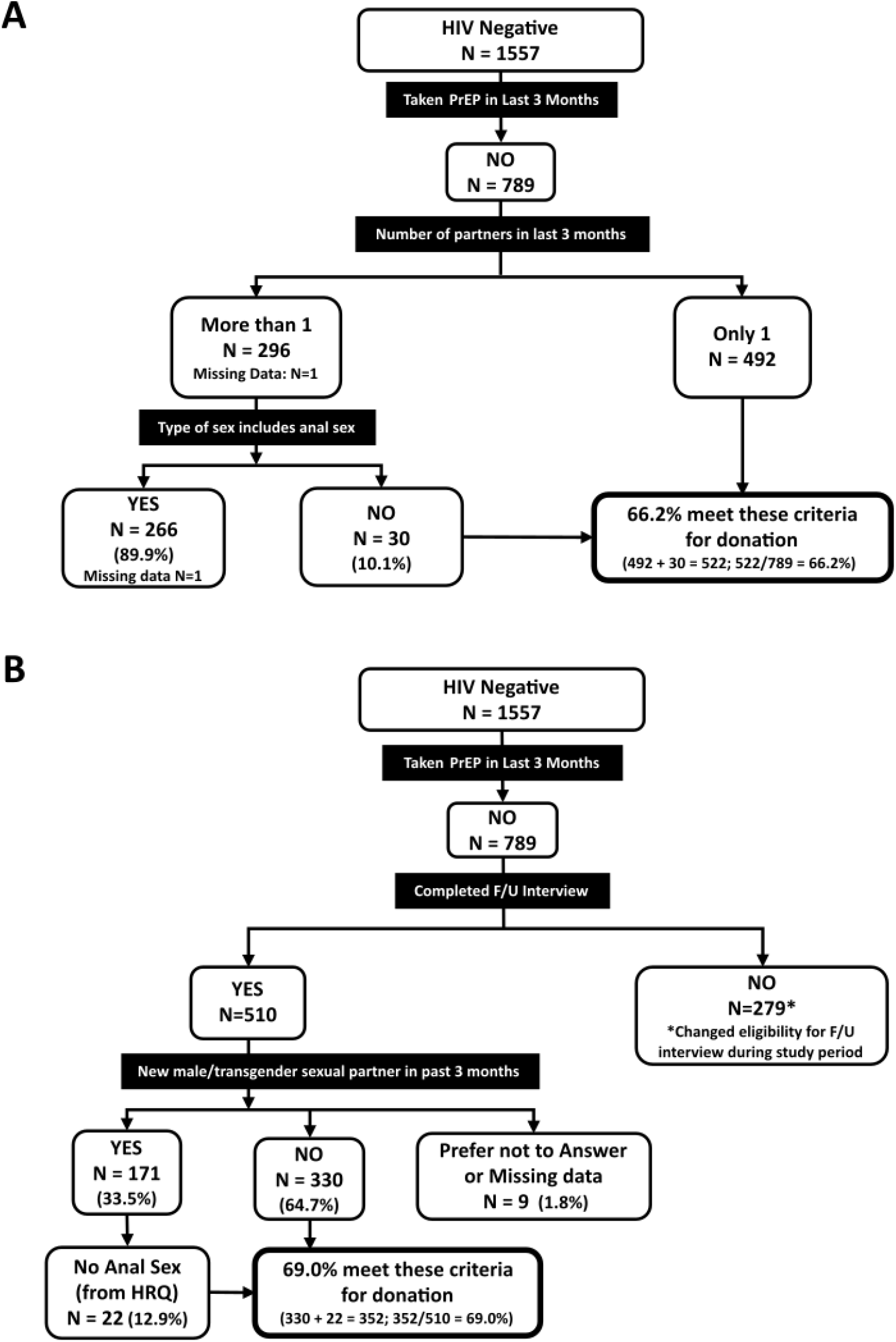
Projected donor eligibility using ADVANCE study results under 2023 draft FDA guidance, based on screening questions regarding (A) number of sexual partners, and (B) change in sexual partners in the past 3 months. F/U – follow-up, HRQ – HIV risk questionnaire.

## Discussion

To assess behaviors associated with HIV risk, we enrolled sexually active MSM living in 8 metropolitan areas in the U.S. who have interest in blood donation. Testing revealed 4 HIV-positive participants, none with an incident HIV infection. Among HIV-negative participants, the HIV risk questions we evaluated demonstrated differences in the risk exposure profiles of the participants, with those not taking PrEP having lower numbers of sexual partners and a lower prevalence of anal sex.

The proportion of HIV-positive MSM was about 1%, consisting of 4 persons with HIV enrolled in this study and 15 individuals reported having HIV during eligibility screening. This percentage is notably lower than surveillance estimates for the overall MSM population in the U.S.^27^ The lower proportion of HIV-positive MSM in this study could reflect knowledge among the participants that HIV-positive persons are not eligible to be blood donors. It could also represent self-selection based on communications among potential participants regarding the study eligibility criteria.

Nearly half of the study population reported recent PrEP use, a proportion higher than reported for other studies of MSM in the U.S.^12,28^ In addition, high levels of educational achievement and income among study participants may indicate higher levels of access to PrEP than for all MSM. The substantial frequency of PrEP use in the study population may have directly contributed to the observed low prevalence of HIV infection. The availability of PrEP and the success of public health initiatives to expand access are critical components to reduce the burden of HIV infection as part of EHE. Those with higher numbers of sexual partners were far more likely to take PrEP than those with fewer partners.

Our study has limitations. First, ADVANCE was not powered to determine if an individual risk-based assessment was as effective at reducing HIV risk as the time-based MSM deferral in donor selection. Second, the study was designed to assess behaviors among sexually active MSM. Therefore, the results of the study are not inclusive of MSM who have not had sex with a male in the past 3 months. If these MSM are not taking PrEP, are HIV-negative and have no other risks, they are eligible to donate under the policy adopted in April 2020.

The number of sexual partners and types of sex reported by study participants are likely generalizable to the MSM population in other areas of the country. However, access to PrEP varies in by geography and is likely to differ even within metropolitan areas, thereby precluding adjustment for these differences.

The study cities were selected because surveillance data indicated elevated HIV incidence among MSM^29^ allowing ADVANCE to assess behaviors associated with HIV in areas with increased HIV risk.

The COVID-19 pandemic may have led to changes in sexual risk behaviors among MSM^30^ as well as reductions in HIV-testing.^31^ At the start of the study, pandemic-associated reduction in close contacts, including sexual contacts, may have occurred. Later in the study enrollment period, when the largest proportion of participants were enrolled, close contacts probably increased with wide availability of COVID-19 vaccination but may have also decreased compared to previous periods because of the mpox virus outbreak.^32^

### Blood Donation Policy Next Steps

The process for the implementation of changes to the donor history questionnaire requires time. Once FDA finalizes the blood donor eligibility guidance document, blood centers will need time to update procedures, modify computer systems, and train staff, before FDA’s final recommendations can be implemented. Changes to the approach to donor selection are carefully monitored to assess whether the risk to blood recipients has changed using programs such as TTIMS^7,8^, where FDA will continue to track the latest data relevant to blood donation and deferral criteria.

The FDA guidance recommends implementation of question hierarchies to be asked of all potential donors. The questions adopted in Canada are similar, with a minor difference that in Canada the use of oral PrEP results is a 4-month deferral. Questions on PrEP use, number of sexual partners, and anal sex among those with multiple partners or new sex partners in the last 3-months will be asked in sequence and will assess these risks in all donors. Donors who are not taking PrEP and have had 1 or no sex partners, regardless of sex or gender, will not be asked about anal sex.

The results from ADVANCE demonstrate that, among sexually active MSM, there are subgroups who test HIV-negative, have had no new sexual partners and only 1 sexual partner within the last 3 months and are likely at lower risk of HIV infection than those with new or multiple sexual partners. These results support the change from excluding all sexually active MSM from blood donation as a single group to an individual risk assessment that defers those who may have higher HIV risk.

## Data Availability

Data from this study are not openly available. Authors should be contacted to discuss data availability.

## Acknowledgements

We acknowledge and thank all persons who were screened for participation in the study and especially those who completed study visits. This study would not have occurred without the dedication and commitment of all study staff at Vitalant, Stanford Blood Center, OneBlood, and American Red Cross, and the assistance of many different partnering organizations, please see [the study website](http://www.advancestudy.org/). Specifically, we thank all the LGBTQ+ community organizations who partnered with the study investigators: Whitman-Walker Health; The Center Orlando; Project More; Los Angeles LGBT Center; City of Hope/Pride in the City; OutMemphis; Friends For Life; CrescentCare; PFLAG New Orleans; New Orleans LGBT Center; Baton Rouge Pride; Ray Rico Freelance; Atlanta Pride, Pridelines; Sunserve, and the Pride Center at Equality Park. In addition, we acknowledge Dr. Marisabel Rodriguez Messan for graphic design and medical writer, Kelly Stimpert for editorial support.

## Supplement

### Sample preparation

Whole blood samples were shipped to Vitalant Research Institute (VRI) for processing into whole blood and plasma aliquots; repository aliquots were stored. Samples were distributed to the 2 testing labs used for this study.

### Laboratory Testing

#### HIV-1 NAT

HIV-1 NAT used in this study is the transcription mediated amplification assay. The detection probabilities (international units/mL) for HIV-1 NAT are a 50% limit of detection of 4.7 IU/mL (95% fiducial limits 4.0 – 5.3), and a 95% limit of detection of 21.2 IU/mL (95% fiducial limits of 18.2 – 25.7). The limit of detection for this assay is 12 copies/mL. This equates to a phase window period of approximately 10-13 days since infection acquisition.^33,34^

#### HIV Serology

Antibody testing is conducted on the Bio-Rad GS HIV-1/HIV-2 PLUS enzyme immunoassay (EIA). This test uses recombinant proteins and synthetic peptides for the detection of antibodies to HIV-1 (Groups M and O) and/or HIV-2 in human serum or plasma. The GS HIV-1/HIV-2 PLUS O EIA testing was conducted on the automated ORTHO® Summit System (OSS).

#### HIV Limiting Antigen

Limiting Antigen (LAg)-Avidity enzyme immunoassay testing (Sedia Biosciences®, Portland, OR) was conducted at VRI. Among those who tested HIV NAT and serology reactive, the LAg Avidity assay is used to define the infection as recently acquired or long-standing.^26^ The estimate of time of infection is based on antibody maturation kinetics and the assay has been shown to classify recent infection as an infection acquired 4 to 6 months before testing.^35,36^

#### Tenofovir

Testing was conducted at VRI. Tenofovir is 1 of the 2 antiretroviral drugs in oral PrEP medications. Tenofovir is a nucleotide analog reverse transcriptase inhibitor with 2 active forms, tenofovir alafenamide and tenofovir disoproxil fumarate. Both are detected by the whole blood enzyme linked immunosorbent assay (ELISA). We used the research use only (Orasure®, Bethlehem, PA) plate-based ELISA with whole blood as the specimen input type. In brief, if the specimen has no detectable tenofovir it will show maximum intensity whereas if tenofovir is present it will show a reduced signal inversely related to the time since last ingestion of oral PrEP. The assay has a 50% reduction in signal at a lower limit of detection of 5 ng/ml (unpublished data per Orasure). As part of the ADVANCE study protocol, results above 55% are classified as tenofovir detected (PrEP reactive) and below 45% as tenofovir not detected (PrEP non-reactive). Those specimens with between 45 – 55% inhibition were retested and if they remained in the range of 45 – 55% inhibition were considered tenofovir inconclusive.

**Table S1.**
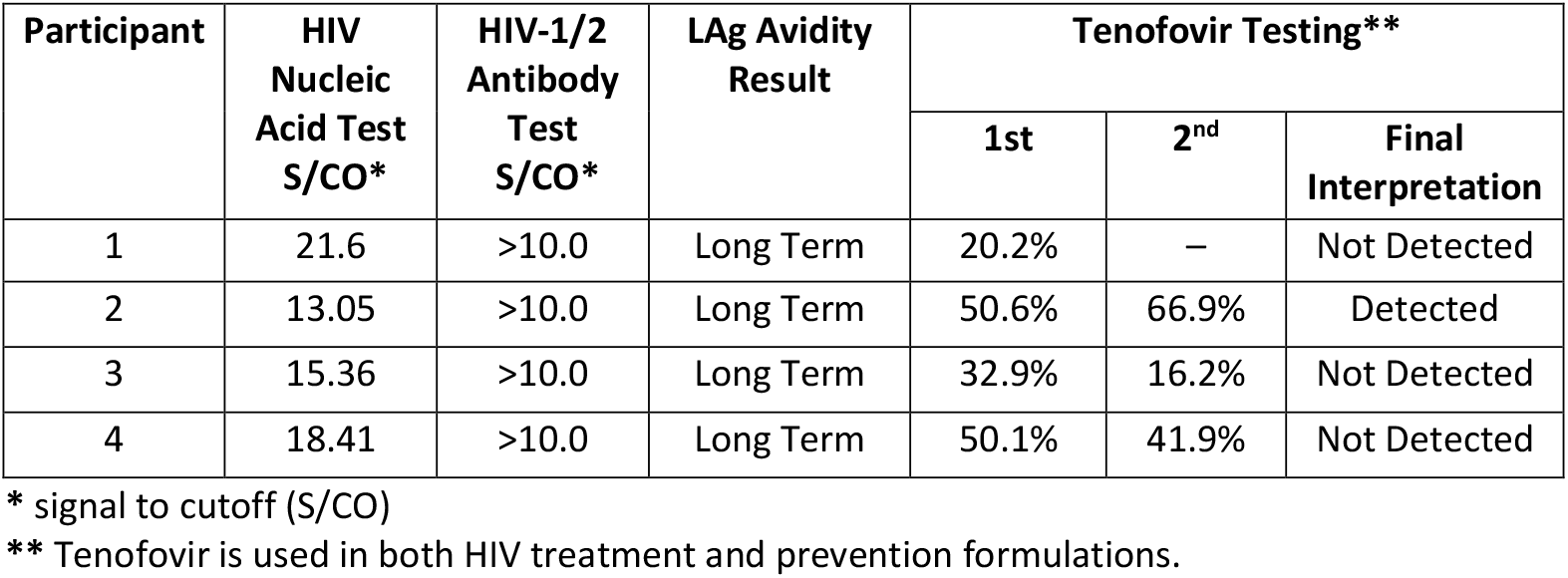
Blood Test Results for Enrolled Study Participants Who Tested HIV-positive.

## Notes

### Competing Interest Statement

The authors have declared no competing interest.

### Funding Statement

This study was funded by the U.S. Food and Drug Administration

### Author Declarations

The Advarra Institutional Review Board (Columbia, MD) gave ethical approval for this work under protocol number 00043278.

